# Ethnic inequalities in age-related patterns of multiple long-term conditions in England: analysis of primary care and nationally representative survey data

**DOI:** 10.1101/2022.08.05.22278462

**Authors:** Brenda Hayanga, Mai Stafford, Catherine L. Saunders, Laia Bécares

## Abstract

**Background:** Having multiple long-term conditions has been associated with a higher treatment burden, reduced quality of life and a higher risk of mortality. Epidemiological evidence suggests that people from minoritised ethnic groups have a higher prevalence of multiple long-term conditions (MLTCs) but questions remain regarding the patterning of MLTCs by age, how this varies for different ethnic group populations, and across conceptualisations of MLTCs (for example, MLTCs with and without mental health conditions). The aim of this study is to examine ethnic inequalities in age-related patterns of MLTCs, and combinations of physical and mental health conditions.

**Methods:** We analysed data from the English GP Patient Survey (GPPS) 2015-2017, and Clinical Practice Research Datalink (CPRD) Aurum from 2016, to give us insight into self-reported and primary care recorded long-term conditions in people aged 18 years and above. We described the association between two or more long-term conditions and age using multilevel regression models adjusting for sex and area-level deprivation with patients nested within GP practices. Similar analyses were repeated for MLTCs that included a mental health condition.

**Findings:** For both self-reported and primary care recorded LTCs, people from most minoritised ethnic groups had a lower prevalence of MLTCs at younger ages compared to their white counterparts. We observed ethnic inequalities from middle age onwards such that in later life, Pakistani, Indian, Black Caribbean and people of Other ethnicity were at an increased risk of having MLTCs compared to white British people. These trends remained after adjusting for area-level deprivation. Compared to white British people, Gypsy and Irish Travellers had higher levels of MLTCs across the age groups, and Chinese people had lower levels. Pakistani and Bangladeshi people aged 50-74 years were more likely than white British people to report two or more LTCs that included a mental health condition. People from other minoritised ethnic groups were less likely to report this compared to white British people.

**Conclusion:** We find clear evidence of ethnic inequalities in MLTCs. It is imperative for health systems to recognise and respond to the higher prevalence of MLTCs that develop by middle age for many minoritised ethnic group people. The lower prevalence of MLTCs that include a mental health condition among some minoritised ethnic group people may be an underestimation due to underdiagnosis and/or inadequate care in primary care and requires further scrutiny.

## INTRODUCTION

### The impact of multiple long-term conditions

It is estimated that approximately 25% of people in the United Kingdom (UK) and approximately 33% of people globally have two or more long-term conditions (MLTCs) ^1-3^. MLTCs pose a number of challenges for patients, health professionals and healthcare systems ^4-7^. For example, many secondary care services are fragmented, highly specialised, and focus on single conditions. Without reform, they are unlikely to meet the needs of patients with MLTCs who require holistic care ^8, 9^. A lack of holistic care for people with MLTCs can complicate treatment, compromise care quality, increase patient dissatisfaction, and further disadvantage patients with MLTCs ^6, 8, 9^. Beyond healthcare, MLTCs are associated with long-term care dependency with people with MLTCs being reported to have an increased risk of becoming care dependant ^10^. A systematic review of studies examining the cost of illness found that the average annual cost of MLTCs ranged from approximately £41 to £204,000_1_ per capita and increased according to the number of MLTCs ^11^. It is clear that having MLTCs has an impact on different facets of society. In the interest of improving the care and quality of life of people with MLTCs, a better understanding of MLTCs is warranted.

### Ethnic inequalities in the prevalence of MLTCs

Over the last decade, there has been a growing number of international studies examining ethnic differences in the prevalence of MLTCs ^12-19^. In the UK, interest in this area is also burgeoning. However, only a handful of studies have counted more than two conditions in their examination of ethnic inequalities in the prevalence of MLTCs ^20-26^. Most of these studies have focused on physical LTCs ^20, 23-26^ and only a few have included mental health conditions in their conceptualisation of MLTCs ^21, 22, 27^. Findings from a systematic review and narrative synthesis of these studies suggest that Black African, Black Caribbean, Indian, Pakistani, Bangladeshi, and people of Black Other, Other Asian, and Mixed ethnicity have a higher prevalence of MLTCs compared to their white British counterparts ^28^. A recent examination of the association between ethnicity and health-related quality of life found that with the exception of Chinese people and Black African men, older people from minoritised ethnic groups reported as many or more LTCs than their white counterparts ^27^. Also, there is evidence to suggest that people with MLTCs from Black African, Black Caribbean, Other Black, Pakistani or Chinese backgrounds have poorer survival compared with white people for total number of conditions or for complex multimorbidity (i.e. MLTCs which involve three or more body systems) ^29^. Thus, not only are there ethnic inequalities in the prevalence of MLTCs, but also in the impact of MLTCs. Whilst some may still consider biological, genetic, behavioural and cultural differences as contributors of ethnic inequalities in multiple conditions, it is widely accepted these factors are not the fundamental cause of ethnic inequalities ^30-33^. Instead, ethnic inequalities in health and by extension, MLTCs, are recognised to result from structural, institutional, and interpersonal racism and discrimination ^34, 35^, which shapes the unequal distribution of resources (e.g. education, employment, income, and housing ^35, 36^) that are strongly associated with poor health.

### The association of MLTCs by age across minoritised ethnic groups

Despite the evidence that MLTCs disproportionately impact many minoritised ethnic group people in the UK, it is less clear how MLTCs are patterned at younger ages as well as mid- and later life for different ethnic group populations. Indeed, some studies report that minoritised ethnic group people develop MLTCs at an earlier age, on average, compared to those from the majority ethnic group ^17, 37^. Others have also found that ethnic inequalities in the prevalence of MLTCs vary at different stages throughout the life course ^12, 13, 15, 19, 38, 39^. For example, Rocca, Boyd (13) examined how the prevalence of MLTCs was patterned by age, sex and ethnicity in a US sample. In their study, the prevalence of two or more conditions among those aged 10-30 years was higher amongst white people compared to Asian and Black people ^13^. From the ages of 31-62 years, Black people had a higher prevalence of two or more conditions compared to all ethnic groups. However, from age 62 years onwards, white people had a higher prevalence of two or more conditions than Black or Asian People ^13^. Asian people had the lowest prevalence of two or more conditions across all age groups with the exception of those aged 75 years and above where their prevalence of two or more conditions was higher than that of Black people but lower when compared to white people ^13^. Whilst such studies provide a nuanced understanding of ethnic inequalities in MLTCs by age, they have been conducted in the US whose minoritised ethnic group populations differ from those in the UK in terms of their migration history, age profile, and nationality. Also, we cannot discount the impact of different healthcare systems on the development and progression of MLTCs given the association between MLTCs and increased healthcare utilisation ^19^. The UK’s healthcare system is markedly different from that of the US as it is publicly funded, with a variety of comprehensive services that are, for the most part, free at the point of use ^40^. It would, therefore, be inappropriate to apply the findings of these studies to the UK context.

Thus far, we have considered ethnic inequalities in the prevalence of MLTCs without making a distinction between MLTCs that consist of only physical health conditions and those that include a mental health condition. However, it is important to make this distinction because over 50% of people who develop a second long-term condition struggle with their mental health as a result of the complex interaction between social, economic and emotional pressures of living with ill health ^36^. In fact, studies show a higher prevalence of mental health conditions, notably depression, in people with long-term physical conditions ^41, 42^. Additionally, having MLTCs that include a mental health condition is associated with a higher symptom burden and functional impairment, greater costs and excess mortality ^7, 38^. Age-related inequalities in MLTCS that include a mental health condition have been reported by Bobo, Yawn (38) who conducted an exploration of the prevalence of combined physical and mental health multimorbidity in a US sample. They found ethnic inequalities in the prevalence of general multimorbidity and multimorbidity that includes a mental health condition; the former increased with age in a linear fashion whilst the latter increased with age with an apparent plateau between ages 50 years and 79 years ^38^. In the UK, ethnic inequalities in the combination of physical and mental health conditions at different ages has not been examined. Findings from such analyses could allow for the identification of populations with a higher prevalence of particular types of MLTCs which, in turn, can facilitate the development of integrated comprehensive services to assist prevention, improve care in the earliest stage of illness, make disease management more efficient in the long-term, and reduce disease burden and progression ^14^.

### Ethnicity data quality

For researchers to fully understand how ethnicity relates to long-term conditions, good quality data is essential ^43^. In the UK, ethnicity recording in routine electronic healthcare records has improved over time but concerns remain about the accuracy and completeness of the data being captured ^43-46^. Recent analysis of the quality of ethnicity coding in hospital datasets by Scobie and colleagues highlighted several data quality problems (e.g. incomplete and inconsistent use of ethnicity codes, excessive use of ‘unknown’, ‘other’ and ‘not stated’ categories)^45^. Crucially, these data quality problems were found to disproportionately affect records for minoritised ethnic group people ^45^. The use of such data can, therefore, introduce bias in the results and/or impede reliable analysis of ethnic inequalities ^45, 46^. Moreover, there is a risk of misclassification brought about by the use of surname recognition software and inconsistencies have been reported when multiple ethnicity coding hierarchies in primary care are used ^47^. To reduce discrepancies and biases in the data, the use of self-reported ethnicity using official classifications of ethnicity is recommended ^44, 46^.

The aim of this study is to use nationally representative data to provide an up-to-date description of how MLTCs vary across ethnic groups in the UK. We describe the pattern of MLTCs, and MLTCs that include a mental health condition, with age across ethnic groups using data from a survey of patients in primary care and routine health data. By using two data sources in this study, we hope to identify potential bias introduced by the aforementioned data quality issues.

## METHODS

### Data

This study uses data from the English GP Patient Survey (GPPS) and Clinical Practice Research Datalink (CPRD) Aurum. The GPPS is a large-scale postal survey which captures reported patient experience of primary care ^48^. It is the only patient experience survey that is standardised across the country and is sent out to over two million people asking them to respond to questions on demographics, their health and their experiences of the healthcare staff and primary care ^49^. The GPPS collects high quality ethnicity data which makes it possible to disaggregate the white ethnic group, thereby allowing for the assessment of patterns of MLTCs by age for marginalised white populations such as the Gypsy, Roma and Traveller community whose health outcomes have been shown to differ markedly from the white British ethnic group ^50^. For this analysis, GPPS data were shared with the University of Cambridge under a data sharing agreement with NHS England. CPRD Aurum contains longitudinal, routinely-collected electronic health records from primary care practices based in England ^51^. The study was reviewed for ethical and methods content and approved by the CPRD team (eRAP protocol number 21_000333). The database captures demographic characteristics, diagnoses and symptoms, prescriptions, vaccination history, laboratory tests, and referrals to hospital and specialist care. As of March 2022, CPRD Aurum had approximately 41 million research acceptable patients ^51^. Full details about the use of data from GPPS and CPRD are available from the respective websites ^52, 53^.

### Study population

For self-reported MLTCs, we combined GPPS data from 2015-6 and 2017 (fieldwork waves January-March 2016, July-September 2016, and January-March 2017) ^54, 55^. In 2017, there were two reminders sent to participants and the overall response rate was 37.5%. For the purpose of this study we included GPPS respondents aged 18 years and above. For primary care reported MLTCs, we used CPRD Aurum data from a random sample of 690,000 patients to achieve an expected sample of at least 600,000 with complete ethnicity data. We included patients who, on 1^st^ January 2016, were 18 years and above, registered in a CPRD practice, and eligible for linkage to Hospital Episode Statistics, ONS mortality data and the 2015 Index of Multiple Deprivation (IMD) score ^56^.

### Measures

To identify long-term conditions in GPPS, we used responses to a question asking participants to specify the long-term conditions they had from a list of 15 conditions (Supplementary table 1). We considered all responses including “another long-term condition”. We defined MLTCs as having two or more LTCs and grouped people into those with 0-1 LTC, those with 2+ LTCs, and those with 2+ LTCs that include a mental health condition. In this study, mental health conditions include everyone who endorsed the response option “a long-term mental health problem” as one of these 15 responses and would therefore be expected to include both common mental health disorders (e.g. anxiety, depression) and severe mental illnesses (e.g. bipolar disorder). The prevalence of long-term conditions reported in response to this question have been compared with other nationally representative survey data (Health Survey for England), and found to give reasonable estimates, apart from long-term mental health conditions, where estimates of prevalence are higher than from other sources ^57^. Ethnic identity was self-ascribed. Respondents were asked to select their ethnic group from 18 ethnic categories based on the England and Wales 2011 Census categories ^58, 59^. Women, older people, and those living in more deprived areas were more likely to have missing ethnicity data (Supplementary Table 2). Participants were grouped into seven age categories (i.e. 18-24, 25-34, 35-44, 45-54, 55-64, 65-74, and 75+). We used IMD scores derived from participants’ postcodes linked to the Office of National Statistics aggregated at the Output Area, which we recoded into quintiles ^54, 55^.

Long-term conditions in CPRD Aurum were identified using diagnosis, symptom and therapy data. We adopted the same approach to defining and operationalising MLTCs as in GPPS described above. We counted the total number of conditions present on 1^st^ January 2016 out of a possible 32 physical and mental health conditions (Supplementary Table 3) that have previously been linked to higher risk of premature death, poor functioning and quality of life, and high user of primary care services ^2^ using and adapting published code lists ^60^. Ethnic identity, usually self-ascribed, was obtained from SNOMED codes recorded by the GP or, where that was missing or incomplete, from linked HES (Hospital Episode Statistics) records. Where multiple values of ethnicity have been recorded, we selected the modal value where this was unique, or the most recent value in line with recommendations made by Mathur, Bhaskaran (43). Categories from the England and Wales 2011 Census ^59^ were used in our analysis but we combined white British, white Irish and other white because these separate categories were not available in HES. Ethnicity data was missing for 14.6% of the sample. Men, younger people and those with fewer LTCs were over-represented in those with missing ethnicity data (Supplementary Table 4). CPRD respondents were also grouped into the seven categories described above. Area-level deprivation data, stratified into quintiles, and derived from participant’s postcode of residence, were based on the 2015 Index of Multiple Deprivation classification at lower super output area ^61^.

### Statistical modelling

Preliminary model development was conducted before analysing GPPS data. In preliminary work, we compared weighted and unweighted analyses and assessed the impact of adjusting for deprivation and GP Practice using multilevel models. In our final analysis we combined responses across two years (the 2015/6 and 2017 survey waves). We report descriptive statistics using numbers of GPPS respondents from both years but estimated weighted percentages in descriptive analyses (to account for non-response and sampling) for 2017 only as cross-sectional survey weights cannot be combined over years ^55^.

For the CPRD analyses unweighted descriptive statistics were calculated. Using data from both datasets, in two separate analyses, we modelled the association between log odds of having MLTCs and age using a logistic regression model with patients nested within GP practices and adjusted for sex and deprivation. From these models we estimated adjusted percentages (“recycled predictions”) at the median deprivation quintile and at estimates at an average of the male and female responses. Age categories were interacted with ethnicity to describe differences in the age-related change in number of LTCs in minoritised ethnic groups compared with the white majority. Because the relationship between multimorbidity and ethnicity varies by age ^29, 62^, we only present adjusted analyses from models which include this interaction term.

We also modelled the log odds of having 2+ LTCs including a mental health condition recorded versus having 0-1 LTC by age and ethnicity.

## RESULTS

The characteristics of the GPPS and CPRD Aurum sample are presented in Table 1. We included 1,432,641 participants from GPPS and 589,246 participants from CPRD Aurum. In both datasets, there were more women than men. People from the white majority ethnic group were generally older than people from minoritised ethnic groups. However, the higher proportions of older people appear to be driven by white British (27.7%) and Irish (37.4%) respondents who are older than Gypsy or Irish Travellers (7.7%) and those from Other White back ground (7.8%). In both the GPPS and CPRD sample, most minoritised ethnic group people were overrepresented in the most deprived areas.

**Table 1.** Description of the CPRD and GPPS samples

|  | GPPS |  |  |  |  | CPRD |  |  |  |  |
| --- | --- | --- | --- | --- | --- | --- | --- | --- | --- | --- |
|  | Number of respondents | Age 65+ (weighted %) | Female (weighted %) | Living in the most deprived 20 % of areas (weighted %) | Living with 2+ long-term conditions (weighted %) | Number of respondents | Age 65+ (%) | Female (%) | Living in the most deprived 20 % of areas (%) | Living with 2+ long-term conditions (%) |
| **All included respondents** | 1,432,641 | 23.9 | 50.7 | 20.1 | 24.1 | 589,246 | 20.6 | 52.8 | 19.2 | 26.8 |
| **White** |  |  |  |  |  | 493,837 | 23.0 | 52.9 | 16.8 | 29.0 |
| White British | 1,172,070 | 27.7 | 51.1 | 16.7 | 26.3 |  |  |  |  |  |
| Irish | 14,553 | 37.4 | 48.4 | 18.6 | 29.8 |  |  |  |  |  |
| Gypsy or Irish Traveller | 365 | 7.7 | 40.4 | 35.3 | 26.7 |  |  |  |  |  |
| Any other White background | 69,851 | 7.8 | 52.3 | 27.4 | 12.2 |  |  |  |  |  |
| **Mixed** |  |  |  |  |  | 9,405 | 5.7 | 54.6 | 29.1 | 13.6 |
| White and Black Caribbean | 3,367 | 4.1 | 57.1 | 33.3 | 16.0 |  |  |  |  |  |
| White and Black African | 1,847 | 3.4 | 47.2 | 38.1 | 15.8 |  |  |  |  |  |
| White and Asian | 3,231 | 5.5 | 48.9 | 20.8 | 14.5 |  |  |  |  |  |
| Any other Mixed background | 3,637 | 7.2 | 52.6 | 28.3 | 17.2 |  |  |  |  |  |
| **Asian** |  |  |  |  |  |  |  |  |  |  |
| Indian | 37,643 | 10.0 | 45.1 | 20.8 | 16.0 | 16,999 | 12.0 | 50.3 | 18.1 | 19.6 |
| Pakistani | 21,690 | 5.9 | 44.1 | 51.7 | 16.1 | 10,554 | 7.2 | 49.0 | 40.3 | 18.4 |
| Bangladeshi | 6,955 | 3.5 | 45.2 | 56.3 | 17.4 | 4,271 | 5.7 | 47.0 | 49.6 | 16.5 |
| Chinese | 7,831 | 6.8 | 52.1 | 22.8 | 8.3 | 6,065 | 5.6 | 56.8 | 20.3 | 5.9 |
| Any other Asian background | 20,199 | 8.5 | 49.3 | 27.3 | 15.9 | 11,654 | 8.0 | 53.0 | 20.3 | 13.9 |
| **Black** |  |  |  |  |  |  |  |  |  |  |
| African | 19,922 | 3.7 | 47.9 | 50.5 | 11.8 | 14,052 | 4.9 | 53.2 | 44.1 | 11.8 |
| Caribbean | 12,573 | 16.3 | 57.8 | 42.3 | 25.8 | 7,813 | 18.7 | 53.9 | 42.5 | 13.3 |
| Any other Black background | 5,819 | 11.4 | 50.4 | 48.6 | 22.7 | 3,930 | 4.5 | 52.3 | 42.5 | 13.3 |
| **Other** |  |  |  |  |  | 10,666 | 6.7 | 52.0 | 29.0 | 10.0 |
| Arab | 3,023 | 6.0 | 38.7 | 37.7 | 16.4 |  |  |  |  |  |
| Any other ethnic group | 28,065 | 10.5 | 44.1 | 40.0 | 19.3 |  |  |  |  |  |

The adjusted prevalence of self-reported MLTCs by age and ethnic group is presented in Figure 1. (See Supplementary Table 5 for the full model estimates). For all ethnic groups, the likelihood of MLTCs was greater at older ages compared with younger ages. Among people aged 18-24 years, people of Indian [Odds Ratio (OR): 0.5, 95% Confidence Interval (CI): 0.4 - 0.6], Pakistani [OR: 0.4, 95% CI: 0.3 - 0.5], Bangladeshi [OR: 0.4, 95% CI: 0.3 - 0.6], Chinese [OR: 0.3, 95% CI: 0.1 - 0.5], African [OR: 0.3, 95% CI: 0.2 - 0.5], Other Asian [OR: 0.4, 95% CI: 0.3 - 0.5], and Other ethnicity [OR: 0.5, 95% CI: 0.4 - 0.7] were less likely to have MLTCs than their white British counterparts (Supplementary table 5). The odds of MLTCs steadily increases for all ethnic groups and by middle age most minoritised ethnic group people fare worse than people with a white British ethnic background. This trend continues in later life such that at ages 75 years and above, the odds of having MLTCS are higher for people of Indian [OR: 1.3, 95% CI: 1.2 – 1.4], Pakistani [OR: 1.2, 95% CI: 1.1-1.4], Black Caribbean [OR: 1.2, 95% CI: 1.1 – 1.3] and Other ethnicity [OR: 1.1, 95% CI: 1.0 – 1.1] (Supplementary table 5). Chinese people are less likely to have 2+ LTCs across all age groups when compared to white British people. In contrast, Gypsy and Irish Travellers aged between 25 and 74 years of age have the highest odds of having 2+ LTCs among all ethnic groups. In sensitivity analysis, we assessed the association between 2+ LTCS and ethnicity with and without adjustment for area-level deprivation and found very little difference between the models (Supplementary Table 6). The age-related patterns of MLTCs by ethnicity in the CPRD sample were very similar (see Supplementary 7 and 8 for CPRD model estimates with and without adjustment for deprivation).

**Figure 1.**
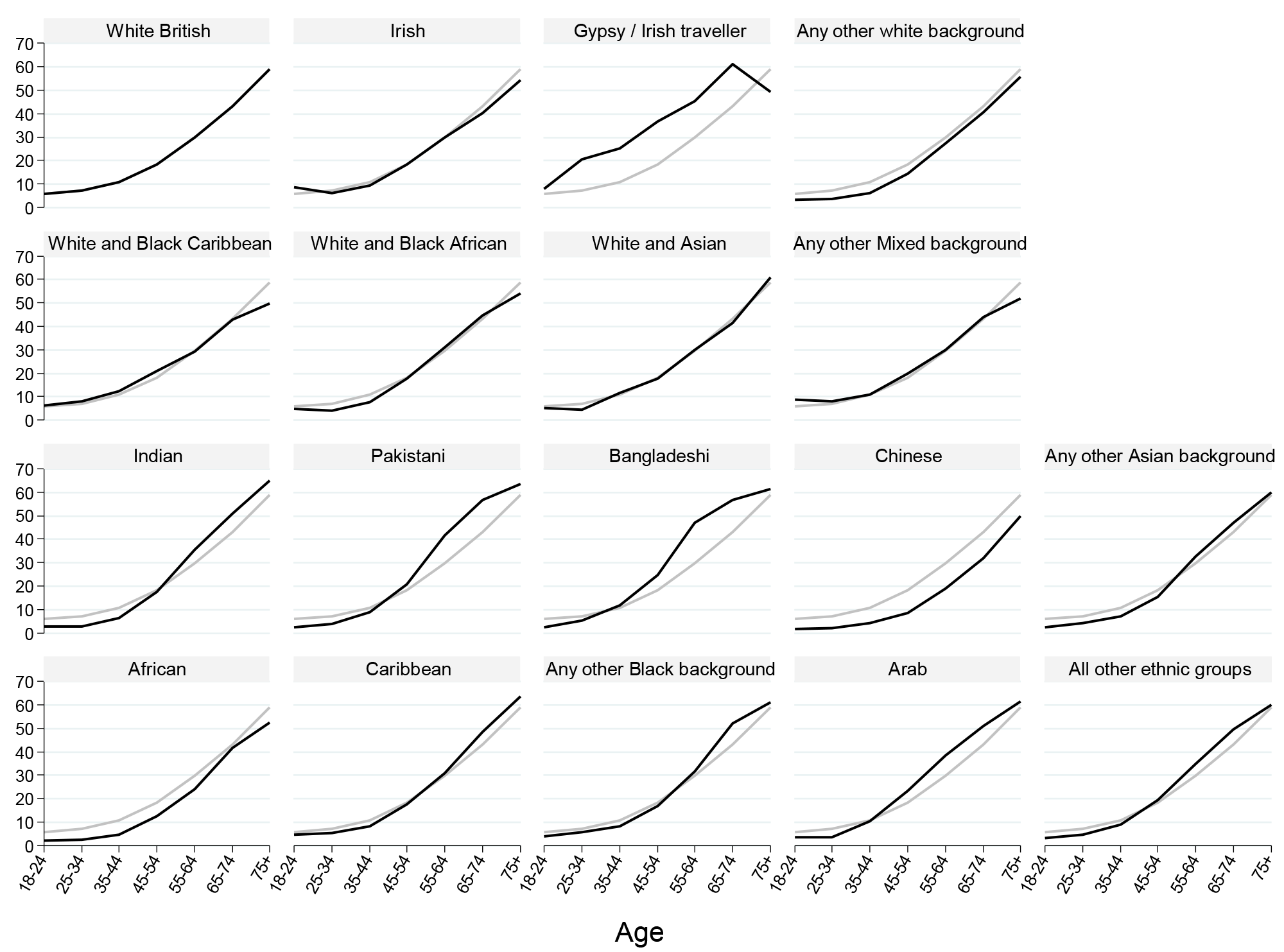
Adjusted percentage of people living with 2+ LTCs, stratified by age and ethnicity from the GPPS sample Note: Estimates for white ethnic group represented in grey for comparison;

Table 2 shows the unadjusted percentages of people living with long-term mental health conditions, and those living with two or more long term conditions including a long-term mental health condition. Compared to the white British ethnic group, the proportion of people living with a self-reported long-term mental health condition is lower for minoritised ethnic group people with the exception of Gypsy or Irish Travellers (14.9%). Similarly, levels of primary care recorded long term mental health conditions are higher in white people than in people from minoritised ethnic groups. When we consider 2+ LTCs including a mental health condition in the CPRD sample, all minoritised ethnic groups have a lower prevalence than their white counterparts. We observe a similar trend in the GPPS sample, however, Gypsy or Irish Travellers, White and Black Caribbean, White and Black African, White and Asian and Other mixed have higher rates of MLTCs (9.9%, 6.1%, 4.4%, 4.6%, 4.8% respectively) that include a mental health condition than the white British ethnic group (3.8%).

**Table 2.** Percentage of people living with long term mental health conditions, and two or more long term conditions including a long term mental health condition. GP Patient Survey and CPRD Aurum

|  |  | GPPS |  | CPRD |  |
| --- | --- | --- | --- | --- | --- |
|  |  | Living with a long-term mental health condition<br>(number, weighted %) | Living with 2+ long-term conditions including a mental health condition<br>(number, weighted %) | Living with a long-term mental health condition<br>(number, %) | Living with 2+ long-term conditions including a mental health condition<br>(number, %) |
| **All included respondents** |  |  |  |  |  |
| **White** |  |  |  | 57099 (11.6) | 41812 (8.5) |
|  | White British | 59624 (6.0) | 39274 (3.8) |  |  |
|  | Irish | 720 (5.9) | 533 (4.3) |  |  |
|  | Gypsy or Irish Traveller | 55 (14.9) | 45 (9.9) |  |  |
|  | Any other White background | 2402 (3.1) | 1515 (1.9) |  |  |
| **Mixed** |  |  |  | 760 (8.1) | 438 (4.7) |
|  | White and Black Caribbean | 292 (9) | 175 (6.1) |  |  |
|  | White and Black African | 124 (7.2) | 69 (4.4) |  |  |
|  | White and Asian | 220 (7.9) | 120 (4.6) |  |  |
|  | Any other Mixed background | 250 (7.4) | 147 (4.8) |  |  |
| **Asian** |  |  |  |  |  |
|  | Indian | 858 (2.1) | 604 (1.4) | 860 (5.1) | 601 (3.5) |
|  | Pakistani | 722 (3) | 518 (1.9) | 672 (6.4) | 476 (4.5) |
|  | Bangladeshi | 269 (3.7) | 192 (2.5) | 281 (6.6) | 190 (4.4) |
|  | Chinese | 151 (2.2) | 68 (0.9) | 161 (2.7) | 57 (0.9) |
|  | Any other Asian background | 571 (3.2) | 383 (1.9) | 518 (4.4) | 331 (2.8) |
| **Black** |  |  |  |  |  |
|  | African | 430 (2.1) | 252 (1.4) | 614 (4.4) | 330 (2.3) |
|  | Caribbean | 481 (4.5) | 327 (2.9) | 596 (7.6) | 436 (5.6) |
|  | Any other Black background | 292 (5.3) | 185 (3.2) | 239 (6.1) | 143 (3.6) |
| **Other** |  |  |  | 597 (5.6) | 330 (3.1) |
|  | Arab | 148 (5.0) | 106 (3.8) |  |  |
|  | Any other ethnic group | 1330 (4.7) | 963 (3) |  |  |

Figure 2 illustrates the adjusted percentages of people living with 2+ LTCs that includes a long-term mental health condition. We present CPRD Aurum data for these analyses because the definition of mental health conditions used in primary care is more comprehensive compared with survey data. (See supplementary Table 9 for full model estimates). Among all ethnic groups, the prevalence of having 2+ LTCs that includes a mental health condition is considerably lower than the prevalence of having 2+ LTCs (note the different scale for Figures 1 and 2). From approximately age 50 years until 74 years, Pakistani and Bangladeshi ethnic group people not only have higher rates of 2+ LTCs that include a mental health condition, but they also have steeper age-related increases when compared to white British ethnic group people. None of the other minoritised ethnic groups had higher rates of 2+ LTCs that include a mental health condition in comparison people of white British ethnicity. These patterns were similar in GPPS data with most minoritised ethnic group people reporting lower prevalence of MLTCs that include a mental health condition when compared to people of white British ethnicity across the age groups (Supplementary Table 10).

**Figure 2.**
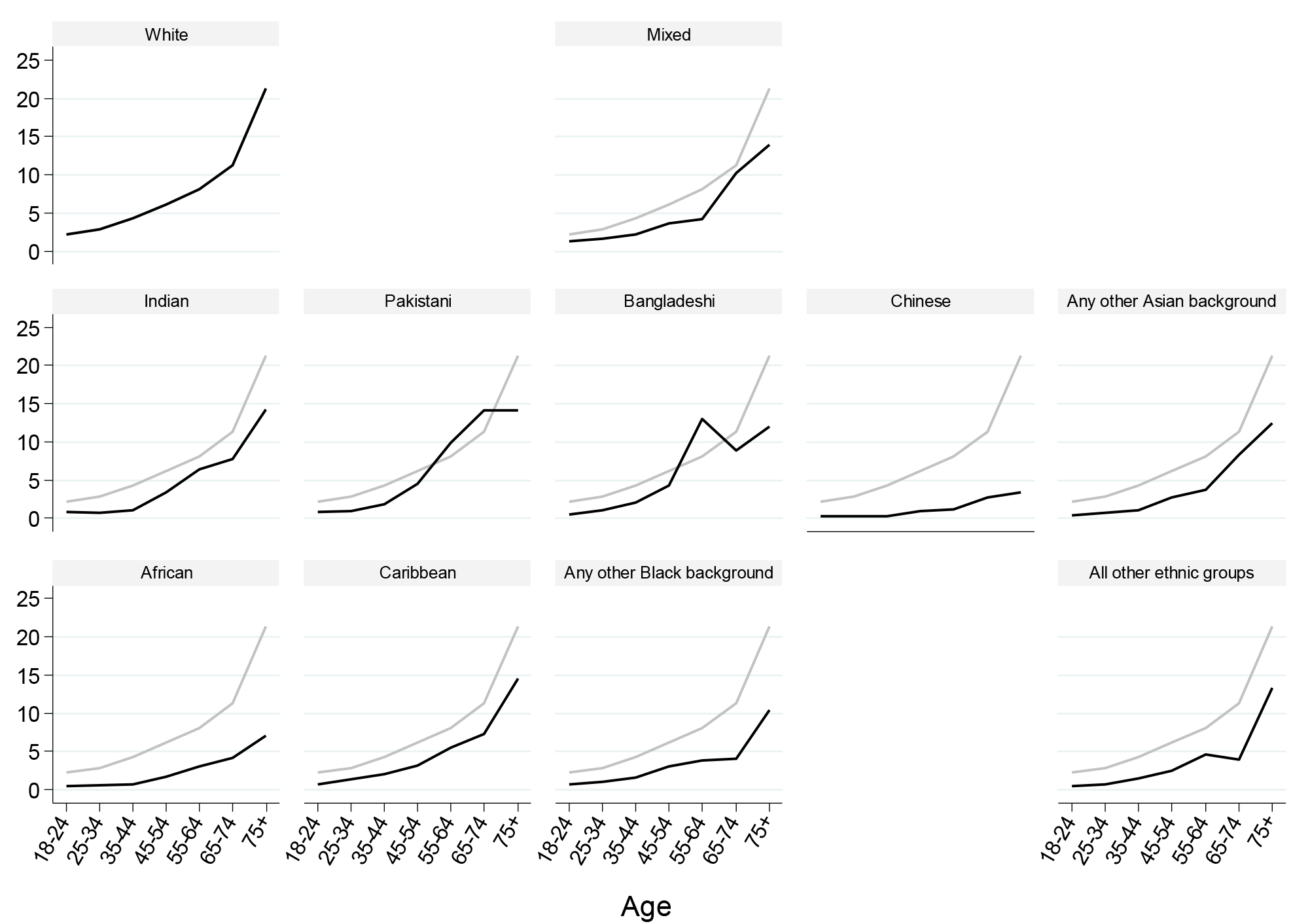
Adjusted percentage of people living with two or more long term conditions including a long term mental health condition. CPRD Aurum sample Note: Estimates for white ethnic group represented in grey for comparison

## DISCUSSION

### Summary of principle findings

In this study, we set out to describe ethnic inequalities in the pattern of MLTCs, and MLTCs that include a mental health condition by age using routine health data from primary care records and self-reported data from a survey of patients in primary care. We found that ethnic inequalities in MLTCs emerge by middle age as most minoritised ethnic group people age 45 years and above had higher prevalence of MLTCs than people of white British ethnicity. These ethnic inequalities extended into later life as older Indian, Pakistani, Black Caribbean and people of Other ethnicity were found to be at an increased risk of MLTCs when compared to older white British people. We also found stark differences between some minoritised ethnic groups. For example, at all age groups, the prevalence of MLTCs was lower among Chinese ethnic group people compared to white British people. In contrast, with the exception of the youngest and oldest age group, the prevalence of MLTCs was higher for Gypsy and Irish Travellers at all age groups. The prevalence of two or more LTCs including a mental health condition was considerably lower than the prevalence of having two or more LTCs. Compared to white people, Pakistani and Bangladeshi people aged between approximately 50 years and 74 years had higher rates of long-term conditions that include a mental health condition. None of the other ethnic groups had higher levels of this type of MLTCs when compared to their white counterparts.

### Comparison with existing literature

Some of the patterns observed in our study are consistent with the general patterns of health inequalities observed by Stopforth and colleagues who found stark and significant ethnic inequalities in limiting long-term illness and self rated health in later life ^63^. These ethnic inequalities were not only greater at older ages, but they persisted over time. In particular, older Black Caribbean, Indian, Pakistani and Bangladeshi people were at increased risk of poor health ^63^. Our findings are also consistent with findings from studies that have specifically explored ethnic inequalities in MLTCs. For example, Watkinson, Sutton (27) examined the association between ethnicity, gender and MLTCs among older adults in the UK. They found that with the exception of Black African men and Chinese men and women, older adults from minoritised ethnic groups reported as many or more long-term conditions as those from the white British majority group ^27^. Their study also found that inequalities were widest for Gypsy or Irish Travellers, Pakistani and Bangladeshi women ^27^. Whilst our study did not compare inequalities between men and women, we found that these three populations were indeed at an increased risk of MLTCs. Our study contributes new knowledge by looking beyond the older population. Such analyses allow for the identification of ethnic group populations that are at particular risk of developing MLTCs at particular stages in the life course. Ultimately, the findings can inform policy makers and practitioners in the development of tailored interventions to address MLTCs.

The findings of this study are also concordant with international studies that have examined age-related ethnic inequalities in MLTCs ^13, 15, 19^. As with our study, these studies also found that ethnic inequalities were not evident at younger ages but developed and widened in mid-life ^13, 15, 19^. In addition, they found that Asian ethnic group people, reported fewer LTCs at most age groups compared to their White, Black or Mixed counterparts ^13, 15, 19^. This finding partially mirrors that of our study in that people of Chinese ethnicity had a lower prevalence of MLTCs than people of White ethnicity across all age groups. Noteworthy is that unlike in our study, the Asian ethnic group in these studies was not disaggregated, thereby, masking ethnic inequalities within individual ethnic groups. The data we used in our study allowed for the disaggregation of not only the Asian ethnic group, but also other ethnic groups, for example, the White ethnic group which consisted of Gypsy and Irish Traveller people who had a higher prevalence of MLTCs compared to their White British counterparts across most age groups.

Our findings concerning ethnic inequalities in MLTCs that involve a mental health condition also partially support those of Bobo and colleagues who explored how the prevalence and patterns of somatic-mental health multimorbidity varied by age, sex and race in a US sample ^38^. They also found that the prevalence of somatic-mental health multimorbidity was lower compared to the prevalence of general multimorbidity. The presence of somatic-mental health multimorbidity was lower in Asian people than in White and Black people ^38^. Just as with the aforementioned international studies, people of Asian ethnicity were aggregated into one broad ethnic group leaving us with a partial understanding of ethnic inequalities in the prevalence of this type of MLTCs. Our study makes an important contribution to this body of literature by disaggregating the major ethnic groups and identifying minoritised ethnic group populations of Asian ethnicity at risk of MLTCs that include a mental health condition (i.e. middle aged Pakistani and Bangladeshi people).

A recent analysis of ethnic inequalities in physical health MLTCs among people with psychosis living in the UK found that Black people were at higher risk of psychosis compared to white people ^64^. The differences between our findings and those of de Freitas and colleagues could be attributed to the fact that the de Freitas study included people under 18 years (age range 13 to 65) and focused on people with a particular type of severe mental illness: psychosis. Others have also found ethnic inequalities in MLTCs that include mental health conditions when they have focused on people with a particular physical condition. For example, Das-Munshi and colleagues found ethnic inequalities not only among people with diabetes, but also among people with diabetes and severe mental illness ^65^. Taken together, our findings and those of other studies illustrate the complexity of ethnic inequalities in health, and raise several questions concerning the underlying processes that lead to differential health outcomes for different minoritised ethnic groups. It is to this that we turn to in the following section.

### Possible mechanisms

The link between socioeconomic status, ethnicity and MLTCs has been articulated by many ^17, 19, 27, 36, 66^. It is acknowledged that people from minoritised ethnic groups are disadvantaged in terms of socioeconomic position, as a result of racism and racial discrimination, that reduce their access and opportunities within employment, education, healthcare, housing, and other sectors ^36^. These negative social outcomes may lead to the development of long-term conditions through lower access to healthcare, inadequate living standards, exposure to environmental stressors and pollutants, reduced levels of health literacy and increased risk of unhealthier behaviours (e.g. physical inactivity, substance abuse, poor diet) ^19, 67^. Given underlying ethnic inequalities in economic and social factors, we adjusted for area-level deprivation (individual-level measures of socioeconomic deprivation were not present in the data analysed). Whilst we acknowledge the limitations of adjusting for socioeconomic status at a single point in time, the fact that ethnic inequalities in MLTCs were still evident suggests that we need to look beyond socioeconomic disadvantage towards the root causes of this disadvantage.

The role of structural racism as the fundamental cause of ethnic inequalities in health has often been overlooked in academic and political realms in favour of more proximal socioeconomic factors ^68^. Bécares and colleagues highlight the role of structural and institutional racism in patterning the unequal distribution of access, power and opportunities (privileging some whilst disadvantaging others) resulting in ethnic inequalities across different domains of society, including socioeconomic position ^68^. As such, racism, in all its forms should be considered in the theorising of, and when available measuring of future analyses of ethnic inequalities in health and their underlying mechanisms.

It is also important to draw on the cumulative advantage/disadvantage theory which has provided a framework for researchers to understand ethnic inequalities in MLTCs among minoritised ethnic group people ^12, 27, 37^. The theory emphasises how early advantage or disadvantage is critical to how cohorts become differentiated over time and recognises the power of social processes and forces that influence the distribution of opportunities among individuals which impact on short-term and long-term outcomes ^69, 70^. When ethnic inequalities in MLTCs are viewed through this lens, the higher prevalence of MLTCs in some minoritised ethnic groups can be attributed to earlier and longer cumulative exposure to risk factors common to many long-term conditions ^37^. In fact, risk factors that affect multiple body systems (e.g. obesity, persistently raised levels of stress and systemic inflammatory markers) are prevalent among minoritised ethnic groups and can lead to earlier development of MLTCs ^37^.

Geronimus and colleagues offer support for this notion ^71^. They examined whether Black people living in the US experienced earlier health deterioration than their white counterparts and found racial differences in the cumulative wear and tear of the body caused by repeated adaptation to stressors ^71^. For them, the challenges that come with living in a race-conscious society which stigmatises and disadvantages Black people may result in disproportionate physiological deterioration, such that a Black person may exhibit the morbidity and mortality typical of a white person who is significantly older ^71^. Evidence from the UK also shows that the detrimental impacts of the experience of racism on health start early in the life course and accumulate over time ^34, 72^. For these reasons, a discussion of processes underlying ethnic inequalities in MLTCs cannot be complete without a consideration of how these upstream forces (e.g. racism, discrimination) intersect with individual level processes ^36, 73, 74^ to shape the health outcomes of different ethnic groups at different stages in life.

Our study finds that ethnic inequalities in the prevalence of MLTCs emerge in middle age and continue into later life for many minoritised ethnic group people. It is possible that this trend could be the result of particular long-term conditions. Mental health conditions tend to increase in prevalence to midlife ^75^. Our analysis shows that based on primary care records, mental health conditions are more prevalent in white people than in minoritised ethnic group people. Arguably, mental health conditions could be driving the higher prevalence of MLTCs in younger white adults. Similarly, diabetes prevalence increases with age and is markedly higher in some minoritised ethnic group people than white people ^76^. It is, therefore, possible that diabetes could be driving the higher prevalence of MLTCs that we observed in mid and later life for some minoritised ethnic group people.

Given that ethnic inequalities in the prevalence of mental health conditions have been reported by several studies ^73, 77-79^, the lower levels of MLTCs that include a mental health condition among most minoritised ethnic group people seen in our study requires further scrutiny. We may be observing an underestimation of the levels of MLTCs that include a mental health condition among some minoritised ethnic group people in primary care. As such, these findings must be interpreted with caution. This underestimation may stem from lower levels of healthcare utilisation caused by language barriers, shame, stigma, mistrust and fear due to racism and discrimination ^80^, which might hinder some minoritised ethnic group people from seeking support for mental health conditions from primary care practitioners. These underestimations may also be the result of misdiagnosis or the failure to detect symptoms by primary healthcare practitioners when minoritised ethnic group people engage with primary care ^81^. Support for this notion is provided by Bignall and colleagues who argue that minoritised ethnic group people are often less likely to access mental health support services through primary care (e.g. via General Practitioners) and are more likely to end up in crisis care ^82^. Evidently, the reasons underlying any underestimations of the prevalence of MLTCs that include a mental health condition among minoritised ethnic groups are likely to involve the complex interaction of several processes.

### Strengths and limitations

A shortcoming of this study is that we assessed MLTCs cross-sectionally and as such, we are unable to draw conclusions about the temporal nature of MLTCs across ethnic group populations. Also, whilst we use a commonly used definition of MLTCs ^2, 3^, this definition fails to capture the severity and duration of the primary care reported and self reported LTCs and their impact on quality of life. Further, it is important to remember that the CPRD (and to a lesser but still important extent GPPS sample) we used in this analysis represent a population that is in contact with healthcare services. Our findings may, therefore, not reflect the true level of inequality as we may have inadvertently excluded those who do not access or utilise health services.

Despite these limitations, this study has several strengths. First, by examining ethnic differences in age-related patterns in LTCs, we provide a nuanced understanding of ethnic inequalities in MLTCs. It is crucial for healthcare systems to recognise and respond to the higher prevalence of MLTCs that develop by middle age for many minoritised ethnic group people. Support with management of MLTCs is required to slow or even halt the progression of MLTCs, thereby, improving the quality of life and well-being for those with MLTCs across different ethnic group populations. At the same time, continued efforts are required across government, public, private sector bodies and the third sector to tackle structural, institutional and interpersonal racism, the fundamental causes of ethnic inequalities in health ^68, 83, 84^.

Second, the use of two data sources, GPPS data and CPRD Aurum meant that we analysed self reported and primary care recorded LTCs. In doing so, we address reporting bias which some consider to be a potential factor driving observed differences in the prevalence of MLTCs between ethnic groups ^85^. Given that similar patterns were observed using self-reported health and primary care records, the inequalities observed in mid- and later life do not seem to be driven by differences in recording. Further, the good quality ethnicity data collected through the GPPS gives us insight into minoritised ethnic groups e.g. White Other, Arab and Gypsy /or Irish Traveler ethnic groups who are often aggregated with other ethnic groups or excluded from analyses due to small numbers ^28^. In doing so, we avoid the essentialisation of minoritised ethnic group people and generate a holistic understanding of ethnic inequalities in the prevalence of MLTCs.

## Conclusion

In this study we examined ethnic inequalities in the age-related patterns of LTCs using self-reported and primary care data. We found ethnic inequalities from midlife onwards for Gypsy or Irish Traveller, older Black Caribbean, Indian, Pakistani and Other ethnic group people, and reported ethnic inequalities in MLTCs that include a mental health condition for Pakistani and Bangladeshi people aged 50-74 years. Whilst the study adds to the sparse body of literature in this field, it raises further questions concerning the accuracy of the prevalence of MLTCs that include a mental health condition in primary care that warrants further investigation. Also, to arrive at a better understanding of age-related inequalities in MLTCs, future work that adopts a longitudinal, intersectional approach is required. Such work would allow for a better understanding of the role that key explanatory factors (including racism and discrimination) play in the development, accumulation and progression of MLTCs over time for different ethnic groups. Analysis that considers how MLTCs impact on quality of life would also be required. It is findings such as these that can help healthcare leads who serve minoritised ethnic group populations to prioritise and tailor clinical management efforts and inform policy makers on the ways in which they can prevent and reduce inequalities in MLTCs at different stages in the life course.

## Data Availability

The study uses individual-level data from General Practice Patient Survey which is available from Ipsos MORI via a data sharing agreement with NHS England. We also use routinely collected individual patient data which can be obtained from Clinical Research Practice Datalink subject to protocol approval via the CPRD Research Data Governance (RDG) Process. Although these data are anonymised, they are considered sensitive data in the UK by the Data Protection Act and, therefore, cannot be shared publicly. Information about applying to use GPPS data can be found at https://gp-patient.co.uk/contact and https://www.cprd.com/data-access respectively

https://gp-patient.co.uk/contact

## DECLARATIONS

### Ethics approval and consent to participate

The study was reviewed for ethical and methods content and approved by the CPRD team (eRAP protocol number 21_000333).

### Consent for publication: Not Applicable

#### Availability of data and materials

The study uses individual-level data from General Practice Patient Survey which is available from Ipsos MORI via a data sharing agreement with NHS England. We also use routinely collected individual patient data which can be obtained from Clinical Research Practice Datalink subject to protocol approval via CPRD’s Research Data Governance (RDG) Process. Although these data are anonymised, they are considered sensitive data in the UK by the Data Protection Act and, therefore, cannot be shared publicly. Information about applying to use data from GPPS and CPRD can be found at https://gp-patient.co.uk/contact and https://www.cprd.com/data-access respectively

### Authors’ contributions

MS, LB and CS conceptualised the study, devised the primary research questions and analysis plan. MS and CS conducted the formal statistical analysis. Output from all analyses was shared with all authors. BH conducted the literature review, drafted the manuscript and revised subsequent drafts. MS, CS and LB critically reviewed, commented and edited the initial and subsequent manuscripts, providing methodological and intellectual feedback. BH submitted the manuscript for publication. All authors have read and agreed to the final version of the submitted manuscript.

## Acknowledgements

This work uses data provided by patients and collected by the NHS as part of their care and support. We thank Jay Hughes at the Health Foundation for his support with the data curation.

## Footnotes

1 Converted from $ 50 to $25000 using exchange rates valid on the 20^th^ of June 2022

