## Supplementary Files for "Ethnic inequalities in age-related patterns of multiple long-term conditions in England: analysis of primary care and nationally representative survey data"

**Supplementary Table 1. Long-term conditions self-reported by participants in the General Practice Patient Survey**

| Mental health conditions | Physical health conditions |
|---|---|
| • Long-term mental health problems | • Alzheimer's disease or dementia<br>• Angina or long-term heart problems<br>• Arthritis or long-term joint problems<br>• Asthma or long-term chest problems<br>• Blindness or severe visual impairment<br>• Cancer in the last five years<br>• Diabetes<br>• Deafness or severe hearing impairment<br>• Epilepsy<br>• High Blood Pressure<br>• Kidney or liver disease<br>• Long-term back problem<br>• Long-term mental health problem<br>• Long-term neurological problem<br>• Another Long-term condition |

**Supplementary Table 2. GPPS participants with complete versus missing ethnicity data**

| | Missing ethnicity | Complete ethnicity | Total n(%) |
|---|---|---|---|
| **Age_groups n(%)** | | | |
| 18-24y | 559(3.3) | 64302(4.0) | 64861(4.0) |
| 25-34y | 1404(8.2) | 148097(9.3) | 149498(9.3) |
| 35-44y | 2130(12.4) | 200124(12.5) | 202254(12.5) |
| 45-54y | 3099(18.1) | 279387(17.5) | 282486(17.5) |
| 55-64y | 3280(19.1) | 320076(20.0) | 323356(20.0) |
| 65-74y | 3492(20.3) | 332418(20.8) | 335910(20.8) |
| 75-84y | 2313(13.5) | 191750(12.0) | 194063(12.0) |
| 85+y | 898(5.2) | 62858(3.9) | 63756(3.9) |
| *Total n(%)* | *17172(100)* | *1599012(100)* | *1616184(100)* |
| **Gender n(%)** | | | |
| Women | 10227(59.2) | 896729 (56.1) | 906956(56.2) |
| Men | 7051(40.8) | 701167(43.9) | 708218(43.9) |
| *Total* | *17278(100)* | *1597896(100)* | *1615174(100)* |
| **IMD n(%)** | | | |
| 1 least deprived | 5348(14.9) | 307014(19.1) | 312362(19.0) |
| 2 | 6124(17.0) | 326620(20.3) | 332744(20.2) |
| 3 | 6767(18.8) | 330818(20.6) | 337585(20.5) |
| 4 | 8219(22.8) | 321969(20.0) | 330188(20.1) |
| 5 | 9566(26.6) | 321408(20.0) | 330974(20.1) |
| *Total* | *36024(100)* | *1607829(100)* | *1643853(100)* |
| **Total LTCs n(%)** | | | |
| No LTCs | 18781(63.4) | 1025348(69.4) | 1044129(69.3) |
| 1+ LTCs | 10855(36.6) | 451351(30.6) | 462206(30.7) |
| Total | *29636(100)* | *1476699(100)* | *1506335(100)* |

**Supplementary Table 3. Long-term conditions counted on 1st January 2016 identified from diagnoses, symptoms and therapy recorded in primary care.**

| **Mental health conditions** | **Physical health conditions** | |
|---|---|---|
| • Alcohol problems<br>• Other psychoactive substance misuse<br>• Anorexia or bulimia<br>• Depression, anxiety & other neurotic, stress-related & somatoform disorders<br>• Schizophrenia (& related non-organic psychosis)/ bipolar disorder | • Asthma (currently treated)<br>• Atrial fibrillation<br>• Blindness and low vision<br>• Bronchiectasis<br>• Chronic kidney disease<br>• Chronic liver disease<br>• Chronic obstructive pulmonary disease (COPD)<br>• Coronary heart disease<br>• Dementia<br>• Diabetes<br>• Diverticulosis | • Multiple sclerosis<br>• New diagnosis of cancer within last 5 years<br>• Parkinson's disease<br>• Peripheral vascular disease<br>• Psoriasis or eczema<br>• Rheumatoid arthritis, other inflammatory polyarthropathies & systematic connective tissue disorders<br>• Stroke & transient ischaemic attack<br>• Thyroid disorders<br>• Viral hepatitis |

**Supplementary Table 4. CPRD patients with complete versus missing ethnicity data**

| | Missing ethnicity | Complete ethnicity |
|---|---|---|
| n | 99585 | 590415 |
| age_baselineg (%) | | |
| 18-29y | 41685 (41.9) | 125853 (21.2) |
| 30-39y | 20857 (20.9) | 115321 (19.5) |
| 40-49y | 14343 (14.4) | 95237 (16.1) |
| 50-59y | 11909 (12.0) | 90677 (15.4) |
| 60-69y | 6752 (6.8) | 68050 (11.5) |
| 70-79y | 2879 (2.9) | 53586 (9.1) |
| 80+y | 1160 (1.2) | 41691 (7.1) |
| gender = women (%) | 41757 (41.9) | 311607 (52.8) |
| IMD2015 | | |
| 1 | 18777 (18.9) | 118930 (20.2) |
| 2 | 19071 (19.2) | 118526 (20.1) |
| 3 | 19038 (19.2) | 114951 (19.5) |
| 4 | 23731 (23.9) | 123581 (21.0) |
| 5 | 18491 (18.7) | 113258 (19.2) |
| Total LTCs; mean (SD) | 0.23 (0.58) | 1.07 (1.46) |

**Supplementary Table 5. Age and ethnicity specific OR for people living with two or more long-term conditions, adjusted for age, gender, deprivation and with an interaction between age and ethnicity. GP Patient Survey**

| | 18-24 | 25-34 | 35-44 | 45-54 | 55-64 | 65-74 | 75+ |
|---|---|---|---|---|---|---|---|
| **White** | | | | | | | |
| White British | reference | reference | reference | reference | reference | reference | reference |
| Irish | 1.5 (0.9 - 2.6) | 0.9 (0.6 - 1.2) | 0.9 (0.7 - 1.0) | 1.0 (0.9 - 1.1) | 1.0 (0.9 - 1.1) | 0.9 (0.8 - 0.9) | 0.8 (0.8 - 0.9) |
| Gypsy or Irish Traveler | 1.4 (0.4 - 4.7) | 3.4 (1.8 - 6.5) | 2.8 (1.6 - 5.0) | 2.6 (1.7 - 4.0) | 2.0 (1.2 - 3.1) | 2.1 (1.1 - 3.8) | 0.7 (0.3 - 1.5) |
| Any other White background | 0.5 (0.4 - 0.6) | 0.5 (0.4 - 0.5) | 0.5 (0.5 - 0.6) | 0.8 (0.7 - 0.8) | 0.9 (0.8 - 0.9) | 0.9 (0.9 - 0.9) | 0.9 (0.8 - 0.9) |
| **Mixed** | | | | | | | |
| White and Black Caribbean | 1.1 (0.8 - 1.5) | 1.1 (0.9 - 1.5) | 1.2 (0.9 - 1.5) | **1.2 (1.0 - 1.4)** | 1.0 (0.8 - 1.2) | 1.0 (0.7 - 1.3) | 0.7 (0.5 - 0.9) |
| White and Black African | 0.8 (0.4 - 1.5) | 0.5 (0.3 - 0.9) | 0.7 (0.5 - 0.9) | 1.0 (0.8 - 1.2) | 1.1 (0.8 - 1.3) | 1.1 (0.7 - 1.5) | 0.8 (0.5 - 1.5) |
| White and Asian | 0.9 (0.6 - 1.3) | 0.6 (0.4 - 0.9) | 1.1 (0.9 - 1.4) | 1.0 (0.8 - 1.2) | 1.0 (0.8 - 1.2) | 0.9 (0.7 - 1.2) | 1.1 (0.8 - 1.6) |
| Any other Mixed background | 1.6 (1.1 - 2.3) | 1.1 (0.9 - 1.5) | 1.0 (0.8 - 1.2) | 1.1 (0.9 - 1.3) | 1.0 (0.8 - 1.2) | 1.0 (0.8 - 1.3) | 0.8 (0.6 - 1.0) |
| **Asian** | | | | | | | |
| Indian | 0.5 (0.4 - 0.6) | 0.4 (0.3 - 0.4) | 0.6 (0.5 - 0.6) | 0.9 (0.9 - 1.0) | **1.3 (1.2 - 1.4)** | **1.4 (1.3 - 1.4)** | **1.3 (1.2 - 1.4)** |
| Pakistani | 0.4 (0.3 - 0.5) | 0.5 (0.4 - 0.6) | 0.8 (0.7 - 0.9) | **1.2 (1.1 - 1.3)** | **1.7 (1.6 - 1.8)** | **1.7 (1.5 - 1.9)** | **1.2 (1.1 - 1.4)** |
| Bangladeshi | 0.4 (0.3 - 0.6) | 0.7 (0.6 - 0.9) | 1.1 (1.0 - 1.3) | **1.5 (1.3 - 1.7)** | **2.1 (1.8 - 2.4)** | **1.7 (1.4 - 2.2)** | 1.1 (0.9 - 1.5) |
| Chinese | 0.3 (0.1 - 0.5) | 0.3 (0.2 - 0.4) | 0.4 (0.3 - 0.5) | 0.4 (0.4 - 0.5) | 0.5 (0.5 - 0.6) | 0.6 (0.5 - 0.7) | 0.7 (0.6 - 0.8) |
| Any other Asian background | 0.4 (0.3 - 0.5) | 0.6 (0.5 - 0.7) | 0.6 (0.6 - 0.7) | 0.8 (0.8 - 0.9) | **1.2 (1.1 - 1.2)** | **1.2 (1.1 - 1.3)** | 1.0 (0.9 - 1.2) |
| **Black** | | | | | | | |
| African | 0.3 (0.2 - 0.5) | 0.3 (0.3 - 0.4) | 0.4 (0.4 - 0.5) | 0.6 (0.6 - 0.7) | 0.8 (0.7 - 0.8) | 0.9 (0.8 - 1.1) | 0.8 (0.6 - 0.9) |
| Caribbean | 0.8 (0.6 - 1.2) | 0.7 (0.6 - 0.9) | 0.7 (0.6 - 0.9) | 1.0 (0.9 - 1.0) | **1.1 (1.0 - 1.1)** | **1.2 (1.1 - 1.4)** | **1.2 (1.1 - 1.3)** |
| Any other Black background | 0.7 (0.4 - 1.2) | 0.8 (0.6 - 1.1) | 0.7 (0.6 - 0.9) | 0.9 (0.8 - 1.0) | **1.1 (1.0 - 1.2)** | **1.4 (1.2 - 1.7)** | 1.1 (0.9 - 1.3) |
| **Other** | | | | | | | |
| Arab | 0.6 (0.3 - 1.2) | 0.5 (0.3 - 0.7) | 1.0 (0.8 - 1.2) | **1.4 (1.2 - 1.6)** | **1.5 (1.2 - 1.8)** | **1.4 (1.1 - 1.8)** | 1.1 (0.8 - 1.6) |
| Any other ethnic group | 0.5 (0.4 - 0.7) | 0.7 (0.6 - 0.8) | 0.8 (0.8 - 0.9) | **1.1 (1.0 - 1.2)** | **1.3 (1.2 - 1.3)** | **1.3 (1.2 - 1.4)** | **1.1 (1.0 - 1.1)** |

**Supplementary Table 6. Sensitivity analysis compared with the model underlying Figure 1. Age and ethnicity specific OR for people living with two or more long-term conditions, adjusted for age and gender and with an interaction between age and ethnicity. GP Patient Survey.**

| | Odds ratio for ethnicity in baseline age group (18-24) | 18-24 | 25-34 | 35-44 | 45-54 | 55-64 | 65-74 | 75+ |
|---|---|---|---|---|---|---|---|---|
| **Odds ratio for age among people with White ethnicity** | | reference | 1.2 (1.2 - 1.3) | 1.9 (1.8 - 2.0) | 3.5 (3.3 - 3.6) | 6.6 (6.3 - 6.9) | 11.5 (11.1 - 12.0) | 21.4 (20.5 - 22.3) |
| | | **Additional age and ethnicity specific OR over and above effect of age and ethnicity in baseline groups** | | | | | | |
| **White** | | | | | | | | |
| White British | reference | reference | reference | reference | reference | reference | reference | reference |
| Irish | 1.5 (0.9 - 2.6) | reference | 0.6 (0.3 - 1.1) | 0.6 (0.3 - 1.0) | 0.7 (0.4 - 1.2) | 0.7 (0.4 - 1.2) | 0.6 (0.4 - 1.1) | 0.6 (0.3 - 1.0) |
| Gypsy or Irish Traveler | 1.7 (0.5 - 5.6) | reference | 2.4 (0.6 - 9.3) | 1.8 (0.5 - 6.9) | 1.7 (0.5 - 6.1) | 1.3 (0.4 - 4.7) | 1.3 (0.3 - 5.1) | 0.4 (0.1 - 1.8) |
| Any other White background | 0.6 (0.5 - 0.7) | reference | 0.9 (0.7 - 1.1) | 1.0 (0.8 - 1.3) | 1.4 (1.2 - 1.8) | 1.6 (1.3 - 2.0) | 1.6 (1.3 - 2.0) | 1.5 (1.3 - 1.9) |
| **Mixed** | | | | | | | | |
| White and Black Caribbean | 1.2 (0.9 - 1.7) | reference | 1.1 (0.7 - 1.6) | 1.1 (0.7 - 1.6) | 1.1 (0.8 - 1.6) | 0.9 (0.6 - 1.3) | 0.9 (0.6 - 1.4) | 0.7 (0.4 - 1.0) |
| White and Black African | 0.9 (0.5 - 1.7) | reference | 0.7 (0.3 - 1.5) | 0.9 (0.5 - 1.8) | 1.3 (0.7 - 2.4) | 1.4 (0.7 - 2.6) | 1.4 (0.7 - 2.9) | 1.2 (0.5 - 2.7) |
| White and Asian | 0.9 (0.6 - 1.4) | reference | 0.7 (0.4 - 1.3) | 1.3 (0.8 - 2.1) | 1.1 (0.7 - 1.8) | 1.2 (0.7 - 1.9) | 1.1 (0.7 - 1.8) | 1.2 (0.7 - 2.2) |
| Any other Mixed background | 1.7 (1.2 - 2.4) | reference | 0.7 (0.5 - 1.1) | 0.6 (0.4 - 1.0) | 0.7 (0.5 - 1.1) | 0.6 (0.4 - 1.0) | 0.7 (0.4 - 1.0) | 0.5 (0.3 - 0.8) |
| **Asian** | | | | | | | | |
| Indian | 0.5 (0.4 - 0.6) | reference | 0.8 (0.6 - 1.0) | 1.2 (0.9 - 1.5) | 2.0 (1.6 - 2.5) | 2.7 (2.1 - 3.4) | 2.9 (2.3 - 3.6) | 2.7 (2.2 - 3.5) |
| Pakistani | 0.5 (0.4 - 0.7) | reference | 1.2 (0.9 - 1.6) | 2.0 (1.5 - 2.5) | 2.9 (2.3 - 3.6) | 4.1 (3.2 - 5.2) | 4.1 (3.2 - 5.3) | 3.0 (2.3 - 3.8) |
| Bangladeshi | 0.5 (0.4 - 0.8) | reference | 1.7 (1.2 - 2.6) | 2.8 (1.9 - 4.0) | 3.6 (2.5 - 5.3) | 5.1 (3.5 - 7.6) | 3.9 (2.6 - 6.0) | 2.8 (1.8 - 4.4) |
| Chinese | 0.3 (0.2 - 0.5) | reference | 1.0 (0.5 - 2.0) | 1.4 (0.7 - 2.6) | 1.5 (0.8 - 2.8) | 1.9 (1.0 - 3.6) | 2.2 (1.2 - 4.1) | 2.6 (1.4 - 4.8) |
| Any other Asian background | 0.4 (0.3 - 0.6) | reference | 1.5 (1.1 - 2.2) | 1.6 (1.2 - 2.3) | 2.1 (1.5 - 2.9) | 2.8 (2.0 - 4.0) | 2.9 (2.1 - 4.0) | 2.6 (1.8 - 3.7) |
| **Black** | | | | | | | | |
| African | 0.4 (0.3 - 0.6) | reference | 0.9 (0.6 - 1.4) | 1.3 (0.9 - 1.8) | 1.9 (1.4 - 2.7) | 2.3 (1.6 - 3.2) | 2.9 (2.0 - 4.1) | 2.3 (1.6 - 3.4) |
| Caribbean | 1.0 (0.7 - 1.5) | reference | 0.9 (0.6 - 1.4) | 0.9 (0.6 - 1.3) | 1.1 (0.8 - 1.7) | 1.3 (0.9 - 1.8) | 1.5 (1.0 - 2.2) | 1.6 (1.1 - 2.3) |
| Any other Black background | 0.9 (0.5 - 1.5) | reference | 1.2 (0.6 - 2.2) | 1.1 (0.6 - 2.0) | 1.3 (0.8 - 2.3) | 1.6 (0.9 - 2.8) | 2.0 (1.1 - 3.5) | 1.6 (0.9 - 2.8) |
| **Other** | | | | | | | | |
| Arab | 0.8 (0.4 - 1.4) | reference | 0.7 (0.4 - 1.5) | 1.6 (0.8 - 3.0) | 2.2 (1.1 - 4.1) | 2.2 (1.2 - 4.2) | 2.0 (1.0 - 3.8) | 1.7 (0.8 - 3.5) |
| Any other ethnic group | 0.6 (0.5 - 0.8) | reference | 1.3 (0.9 - 1.7) | 1.6 (1.2 - 2.1) | 2.0 (1.5 - 2.7) | 2.3 (1.7 - 3.0) | 2.3 (1.7 - 3.0) | 1.9 (1.4 - 2.5) |

**Supplementary Table 7. CPRD version of the model for Supplementary Table 6 underlying Figure 1. Age and ethnicity specific OR for people living with two or more long-term conditions, adjusted for age, gender, deprivation and with an interaction between age and ethnicity**

| | Odds ratio for ethnicity in baseline age group (18-24) | 18-24 | 25-34 | 35-44 | 45-54 | 55-64 | 65-74 | 75+ |
|---|---|---|---|---|---|---|---|---|
| **Odds ratio for age among people with White ethnicity** | | reference | 1.3 (1.3-1.4) | 2.1 (2.0-2.1) | 3.5 (3.4-3.6) | 6.5 (6.3-6.7) | 13.6 (13.2-14.0) | 37.2 (35.9-38.4) |
| | | **Additional age and ethnicity specific OR over and above effect of age and ethnicity in baseline groups** | | | | | | |
| **White** | reference | reference | reference | reference | reference | reference | reference | reference |
| **Mixed** | 0.6 (0.5-0.7) | reference | 0.9 (0.7-1.1) | 0.9 (0.7-1.1) | 1.1 (0.9-1.4) | 1.1 (0.9-1.4) | 1.6 (1.2-2.2) | 1.2 (0.9-1.8) |
| **Asian** | | | | | | | | |
| Indian | 0.6 (0.5-0.7) | reference | 0.6 (0.5-0.8) | 0.9 (0.7-1.1) | 1.7 (1.4-2.2) | 2.5 (2.0-3.1) | 2.5 (2.0-3.1) | 1.6 (1.2-2.1) |
| Pakistani | 0.6 (0.5-0.7) | reference | 0.8 (0.6-1.0) | 1.2 (1.0-1.5) | 2.3 (1.8-2.9) | 3.1 (2.4-4.0) | 3.1 (2.3-4.2) | 1.4 (1.0-1.9) |
| Bangladeshi | 0.5 (0.3-0.6) | reference | 0.9 (0.6-1.3) | 1.7 (1.2-2.5) | 2.7 (1.8-3.9) | 4.4 (2.9-6.6) | 2.8 (1.7-4.5) | 1.5 (0.9-2.6) |
| Chinese | 0.1 (0.1-0.2) | reference | 1.1 (0.6-1.8) | 1.2 (0.7-2.1) | 2.3 (1.4-3.8) | 3.6 (2.3-5.6) | 4.4 (2.7-7.2) | 3.3 (2.0-5.7) |
| Any other Asian background | 0.3 (0.2-0.4) | reference | 0.9 (0.7-1.3) | 1.6 (1.2-2.1) | 2.7 (2.0-3.6) | 3.7 (2.7-5.0) | 3.8 (2.8-5.2) | 2.6 (1.8-3.8) |
| **Black** | | | | | | | | |
| African | 0.2 (0.2-0.3) | reference | 1.2 (0.9-1.6) | 1.4 (1.1-1.9) | 2.6 (2.0-3.4) | 3.5 (2.6-4.7) | 3.4 (2.5-4.7) | 1.8 (1.3-2.7) |
| Caribbean | 0.4 (0.3-0.5) | reference | 1.6 (1.1-2.3) | 1.7 (1.2-2.4) | 2.1 (1.5-2.9) | 2.5 (1.8-3.5) | 2.9 (2.1-4.2) | 2.5 (1.7-3.5) |
| Any other Black background | 0.5 (0.4-0.7) | reference | 1.0 (0.7-1.4) | 0.9 (0.7-1.4) | 1.5 (1.0-2.1) | 1.2 (0.8-1.8) | 0.9 (0.6-1.6) | 1.1 (0.6-1.9) |
| **Other** | 0.2 (0.2-0.3) | reference | 1.0 (0.7-1.5) | 1.5 (1.0-2.0) | 2.0 (1.5-2.8) | 2.7 (2.0-3.8) | 2.4 (1.7-3.4) | 2.8 (1.9-4.1) |

**Supplementary Table 8. CPRD version of the model for Supplementary Table 6. Age and ethnicity specific OR for people living with two or more long-term conditions, adjusted for age, gender, deprivation and with an interaction between age and ethnicity. No adjustment for deprivation.**

| | Odds ratio for ethnicity in baseline age group (18-24) | 18-24 | 25-34 | 35-44 | 45-54 | 55-64 | 65-74 | 75+ |
|---|---|---|---|---|---|---|---|---|
| **Odds ratio for age among people with White ethnicity** | | 1.3 (1.3-1.4) | 2.0 (2.0-2.1) | 3.4 (3.3-3.5) | 6.2 (6.1-6.4) | 12.9 (12.5-13.3) | 35.1 (34.0-36.3) | 1.3 (1.3-1.4) |
| | | **Additional age and ethnicity specific OR over and above effect of age and ethnicity in baseline groups** | | | | | | |
| **White** | reference | reference | reference | reference | reference | reference | reference | reference |
| **Mixed** | 0.6 (0.5-0.8) | reference | 0.9 (0.7-1.1) | 0.9 (0.7-1.1) | 1.1 (0.9-1.4) | 1.1 (0.9-1.5) | 1.6 (1.2-2.2) | 1.3 (0.9-1.8) |
| **Asian** | | | | | | | | |
| Indian | 0.6 (0.5-0.7) | reference | 0.6 (0.5-0.8) | 0.9 (0.7-1.1) | 1.7 (1.4-2.2) | 2.6 (2.1-3.2) | 2.5 (2.0-3.2) | 1.6 (1.3-2.1) |
| Pakistani | 0.6 (0.5-0.7) | reference | 0.8 (0.6-1.0) | 1.2 (1.0-1.6) | 2.3 (1.8-2.9) | 3.2 (2.5-4.1) | 3.2 (2.4-4.3) | 1.4 (1.0-2.0) |
| Bangladeshi | 0.5 (0.4-0.7) | reference | 0.9 (0.6-1.3) | 1.7 (1.2-2.5) | 2.7 (1.8-3.9) | 4.4 (2.9-6.6) | 2.9 (1.8-4.8) | 1.6 (0.9-2.7) |
| Chinese | 0.1 (0.1-0.2) | reference | 1.1 (0.6-1.8) | 1.2 (0.7-2.1) | 2.3 (1.4-3.8) | 3.6 (2.3-5.6) | 4.5 (2.7-7.2) | 3.5 (2.0-5.9) |
| Any other Asian background | 0.3 (0.2-0.4) | reference | 0.9 (0.7-1.3) | 1.6 (1.2-2.1) | 2.7 (2.0-3.7) | 3.7 (2.7-5.0) | 3.8 (2.8-5.3) | 2.6 (1.8-3.8) |
| **Black** | | | | | | | | |
| African | 0.3 (0.2-0.3) | reference | 1.2 (0.9-1.6) | 1.5 (1.1-2.0) | 2.7 (2.0-3.6) | 3.7 (2.8-4.9) | 3.6 (2.6-5.0) | 1.9 (1.3-2.8) |
| Caribbean | 0.4 (0.3-0.6) | reference | 1.6 (1.1-2.3) | 1.7 (1.2-2.5) | 2.1 (1.5-2.9) | 2.6 (1.8-3.6) | 3.0 (2.1-4.3) | 2.5 (1.8-3.6) |
| Any other Black background | 0.6 (0.4-0.8) | reference | 1.0 (0.7-1.4) | 1.0 (0.7-1.4) | 1.5 (1.1-2.1) | 1.3 (0.8-1.8) | 1.0 (0.6-1.6) | 1.1 (0.7-1.9) |
| **Other** | 0.2 (0.2-0.3) | reference | 1.0 (0.7-1.5) | 1.5 (1.1-2.1) | 2.1 (1.5-2.9) | 2.7 (2.0-3.8) | 2.4 (1.7-3.5) | 2.7 (1.9-4.0) |

**Supplementary Table 9. Model for Figure 2. Age and ethnicity specific OR for people living with two or more long term conditions including a long term mental health problem, adjusted for age, gender, deprivation and with an interaction between age and ethnicity. CPRD Aurum sample**

| | Odds ratio for ethnicity in baseline age group (18-24) | 18-24 | 25-34 | 35-44 | 45-54 | 55-64 | 65-74 | 75+ |
|---|---|---|---|---|---|---|---|---|
| **Odds ratio for age among people with White ethnicity** | | reference | 1.3 (1.2-1.4) | 2.0 (1.9-2.1) | 2.9 (2.8-3.0) | 3.9 (3.7-4.1) | 5.6 (5.4-5.9) | 12.0 (11.4-12.6) |
| | | **Additional age and ethnicity specific OR over and above effect of age and ethnicity in baseline groups** | | | | | | |
| **White** | reference | reference | reference | reference | reference | reference | reference | reference |
| **Mixed** | 0.6 (0.5-0.8) | reference | 1.0 (0.7-1.3) | 0.8 (0.6-1.2) | 1.0 (0.7-1.4) | 0.8 (0.6-1.2) | 1.5 (0.9-2.4) | 1.0 (0.6-1.8) |
| **Asian** | | | | | | | | |
| Indian | 0.4 (0.3-0.5) | reference | 0.6 (0.4-0.9) | 0.6 (0.4-0.9) | 1.4 (1.0-2.0) | 2.0 (1.4-2.9) | 1.7 (1.1-2.6) | 1.6 (1.0-2.4) |
| Pakistani | 0.4 (0.3-0.5) | reference | 1.5 (0.8-3.0) | 2.0 (1.0-3.9) | 2.9 (1.5-5.9) | 7.2 (3.6-14.6) | 3.3 (1.4-7.8) | 2.1 (0.8-5.5) |
| Bangladeshi | 0.2 (0.1-0.4) | reference | 0.8 (0.5-1.2) | 1.1 (0.8-1.6) | 2.0 (1.3-2.9) | 3.3 (2.2-4.9) | 3.5 (2.1-5.6) | 1.6 (1.0-2.8) |
| Chinese | 0.1 (0.1-0.2) | reference | 1.1 (0.5-2.5) | 0.7 (0.3-1.8) | 1.5 (0.6-3.5) | 1.4 (0.5-3.4) | 2.3 (0.9-6.0) | 1.3 (0.4-4.3) |
| Any other Asian background | 0.2 (0.1-0.3) | reference | 1.3 (0.8-2.2) | 1.3 (0.8-2.1) | 2.2 (1.3-3.6) | 2.3 (1.4-3.9) | 3.7 (2.2-6.4) | 2.7 (1.4-5.2) |
| **Black** | | | | | | | | |
| African | 0.2 (0.1-0.3) | reference | 1.0 (0.6-1.5) | 0.8 (0.5-1.2) | 1.3 (0.9-2.0) | 1.7 (1.1-2.8) | 1.7 (1.0-3.0) | 1.4 (0.7-2.6) |
| Caribbean | 0.3 (0.2-0.5) | reference | 1.6 (0.9-2.8) | 1.6 (0.9-2.7) | 1.7 (1.0-2.9) | 2.3 (1.3-3.9) | 2.1 (1.2-3.8) | 2.1 (1.2-3.7) |
| Any other Black background | 0.3 (0.2-0.5) | reference | 1.1 (0.6-2.1) | 1.1 (0.6-2.1) | 1.5 (0.9-2.8) | 1.4 (0.7-2.8) | 1.1 (0.4-3.0) | 1.4 (0.5-3.6) |
| **Other** | 0.2 (0.2-0.3) | reference | 1.1 (0.7-1.7) | 1.4 (0.9-2.2) | 1.7 (1.1-2.7) | 2.3 (1.4-3.7) | 1.4 (0.8-2.5) | 2.4 (1.3-4.4) |

**Supplementary Table 10. GPPS version of the model for Supplementary Table 9. Age and ethnicity specific OR for people living with two or more long term conditions including a long term mental health problem, adjusted for age, gender, deprivation and with an interaction between age and ethnicity**

| | **Odds ratio for ethnicity in baseline age group (18-24)** | **18-24** | **25-34** | **35-44** | **45-54** | **55-64** | **65-74** | **75+** |
|---|---|---|---|---|---|---|---|---|
| **Odds ratio for age among people with White ethnicity** | | reference | 1.1 (1.0 - 1.2) | 1.5 (1.4 - 1.6) | 1.8 (1.7 - 1.9) | 1.6 (1.5 - 1.7) | 0.8 (0.7 - 0.8) | 0.5 (0.5 - 0.5) |
| | | **Additional age and ethnicity specific OR over and above effect of age and ethnicity in baseline groups** | | | | | | |
| **White** | | | | | | | | |
| White British | reference | reference | reference | reference | reference | reference | reference | reference |
| Irish | 1.9 (1.0 - 3.7) | reference | 0.4 (0.2 - 0.8) | 0.5 (0.2 - 1.0) | 0.7 (0.3 - 1.3) | 0.6 (0.3 - 1.3) | 0.5 (0.3 - 1.1) | 0.6 (0.3 - 1.2) |
| Gypsy or Irish Traveler | 2.3 (0.3 - 17.4) | reference | 0.8 (0.1 - 6.9) | 1.7 (0.2 - 14.1) | 1.7 (0.2 - 13.5) | 1.0 (0.1 - 8.7) | 1.9 (0.2 - 17.6) | |
| Any other White background | 0.4 (0.3 - 0.5) | reference | 0.7 (0.5 - 1.0) | 0.8 (0.6 - 1.2) | 1.7 (1.2 - 2.3) | 2.4 (1.7 - 3.3) | 2.8 (2.0 - 4.1) | 3.0 (2.0 - 4.4) |
| Mixed | | | | | | | | |
| White and Black Caribbean | 1.2 (0.8 - 1.8) | reference | 0.9 (0.5 - 1.6) | 1.0 (0.6 - 1.7) | 1.1 (0.6 - 1.8) | 0.7 (0.3 - 1.2) | 0.5 (0.1 - 1.6) | 0.9 (0.3 - 2.9) |
| White and Black African | 0.9 (0.4 - 2.0) | reference | 0.4 (0.1 - 1.4) | 0.5 (0.2 - 1.4) | 0.9 (0.4 - 2.3) | 1.2 (0.5 - 3.2) | 0.6 (0.1 - 2.4) | |
| White and Asian | 1.1 (0.6 - 1.9) | reference | 0.6 (0.3 - 1.3) | 1.2 (0.6 - 2.4) | 0.8 (0.4 - 1.5) | 0.8 (0.4 - 1.7) | 0.3 (0.1 - 1.5) | 1.5 (0.4 - 5.4) |
| Any other Mixed background | 0.9 (0.5 - 1.7) | reference | 1.1 (0.5 - 2.3) | 1.1 (0.5 - 2.2) | 1.1 (0.5 - 2.2) | 1.0 (0.5 - 2.2) | 0.8 (0.3 - 2.1) | 1.0 (0.3 - 3.2) |
| Asian | | | | | | | | |
| Indian | 0.2 (0.1 - 0.3) | reference | 1.0 (0.6 - 1.8) | 1.0 (0.6 - 1.7) | 1.9 (1.1 - 3.3) | 2.9 (1.7 - 4.8) | 3.9 (2.3 - 6.7) | 6.8 (3.9 - 11.8) |
| Pakistani | 0.2 (0.1 - 0.3) | reference | 1.2 (0.7 - 2.0) | 1.4 (0.9 - 2.3) | 2.2 (1.4 - 3.5) | 3.1 (2.0 - 4.9) | 5.3 (3.2 - 8.6) | 5.7 (3.3 - 9.9) |
| Bangladeshi | 0.3 (0.1 - 0.5) | reference | 1.3 (0.6 - 2.6) | 1.7 (0.9 - 3.3) | 2.0 (1.0 - 3.8) | 2.4 (1.2 - 4.8) | 5.3 (2.3 - 12.0) | 6.9 (2.9 - 16.5) |
| Chinese | 0.3 (0.1 - 0.7) | reference | 0.4 (0.1 - 1.1) | 0.4 (0.2 - 1.1) | 0.4 (0.2 - 1.2) | 1.0 (0.4 - 2.4) | 1.5 (0.5 - 4.2) | 2.0 (0.5 - 7.0) |
| Any other Asian background | 0.3 (0.2 - 0.5) | reference | 1.5 (0.8 - 2.8) | 1.2 (0.6 - 2.1) | 1.5 (0.8 - 2.7) | 1.7 (1.0 - 3.2) | 2.2 (1.2 - 4.2) | 2.5 (1.1 - 5.6) |
| Black | | | | | | | | |
| African | 0.3 (0.2 - 0.4) | reference | 0.6 (0.3 - 1.2) | 0.6 (0.4 - 1.1) | 0.8 (0.4 - 1.3) | 1.0 (0.5 - 1.8) | 2.0 (1.0 - 4.0) | 1.8 (0.6 - 5.1) |
| Caribbean | 0.4 (0.2 - 0.7) | reference | 1.5 (0.7 - 3.3) | 1.2 (0.5 - 2.5) | 1.3 (0.6 - 2.6) | 1.6 (0.8 - 3.2) | 1.2 (0.5 - 2.7) | 2.1 (1.0 - 4.6) |
| Any other Black background | 0.5 (0.2 - 1.1) | reference | 0.7 (0.2 - 2.1) | 1.2 (0.5 - 3.1) | 1.0 (0.4 - 2.6) | 1.7 (0.7 - 4.3) | 1.3 (0.5 - 3.7) | 1.2 (0.4 - 3.9) |
| Other | | | | | | | | |
| Arab | 0.7 (0.3 - 1.5) | reference | 0.5 (0.2 - 1.5) | 0.9 (0.3 - 2.1) | 1.1 (0.4 - 2.7) | 1.8 (0.7 - 4.5) | 1.4 (0.4 - 4.4) | 0.7 (0.1 - 5.6) |
| Any other ethnic group | 0.4 (0.2 - 0.6) | reference | 1.1 (0.6 - 1.8) | 1.3 (0.8 - 2.1) | 1.9 (1.2 - 3.0) | 2.3 (1.5 - 3.7) | 3.0 (1.9 - 4.9) | 3.7 (2.2 - 6.2) |
